# Unplanned ICU transfer from post-anesthesia care unit following cerebral surgery: A retrospective study

**DOI:** 10.1101/2022.03.13.22272048

**Authors:** Qinqin Cao, Chengjuan Fan, Wei Li, Shuling Bai, Hemin Dong, Haihong Meng

**Author notes:** **Correspondence:** Haihong Meng, mailing address: No. 89 Guhuai Road, Rencheng District, Jining City, Shandong Province, China; E-mai.

## Abstract

**Background:** Unplanned transfer to intensive care unit (ICU) lead to reduced trust of patients and their families in medical staff and challenge medical staff to allocate scarce ICU resources. This study aimed to explore the incidence and risk factors of unplanned transfer to ICU during emergence from general anesthesia after cerebral surgery, and to provide guidelines for preventing unplanned transfer from post-anesthesia care unit (PACU) to ICU following cerebral surgery.

**Methods:** This was a retrospective case-control study and included patients with unplanned transfer from PACU to ICU following cerebral surgery between January 2016 and December 2020. The control group comprised patients matched (2:1) for age (±5 years), sex, and operation date (±48 hours) as those in the case group. Stata14.0 was used for statistical analysis, and p <0.05 indicated statistical significance.

**Results:** A total of 11,807 patients following cerebral surgery operations were cared in PACU during the study period. Of the 11,807 operations, 81 unscheduled ICU transfer occurred (0.686%). Finally, 76 patients were included in the case group, and 152 in the control group. The following factors were identified as independent risk factors for unplanned ICU admission after neurosurgery: low mean blood oxygen (OR=1.57, 95%CI: 1.20–2.04), low mean albumin (OR=1.14, 95%CI: 1.03–1.25), slow mean heart rate (OR=1.04, 95%CI: 1.00–1.08), blood transfusion (OR=2.78, 95%CI: 1.02–7.58), emergency surgery (OR=3.08, 95%CI: 1.07–8.87), lung disease (OR=2.64, 95%CI: 1.06–6.60), and high mean blood glucose (OR=1.71, 95%CI: 1.21–2.41).

**Conclusion:** We identified independent risk factors for unplanned transfer from PACU to ICU after cerebral surgery based on electronic medical records. Early identification of patients who may undergo unplanned ICU transfer after cerebral surgery is important to provide guidance for accurately implementing a patient’s level of care.

## Introduction

The post-anesthesia care unit (PACU) is where surgical patients under general anesthesia receive monitoring and care immediately after surgery, and is an intermediate transitional unit to provide care for patients^[1, 2]^. Postoperative patients were admitted to the PACU and closely cared for by the anesthetic nurse and anesthesiologist until they recovered from the anesthesia effects, and then safely transferred to the ward by the anesthetic nurse^[3]^.

Unplanned transfer of patients from the PACU to the ICU after surgery is one of the common critical events in the PACU^[4, 5]^. It is well-known that there is a shortage of ICU beds worldwide^[6-8]^. Patients without plans to transfer to the ICU before surgery, and their transport time is often delayed compared with those who have reserved ICU beds ^[9]^. Limited ICU resources and delays in transfers are associated with increased hospitalization costs and morbidity, which can negatively affect their treatment and prognosis^[10]^.

The risk of complications in cerebral surgery is high, and patients are prone to unpredictable changes after surgery^[11, 12]^. In previous clinical practice, patients were usually admitted to the ICU for 12–24 hours of observation and treatment after cerebral surgery; most of them were transferred to the general ward the next day^[13-15]^. With the recent advancement in medicine and surgery, many studies have shown that patients do not need to be routinely transferred to the ICU after cerebral surgery; patients should be strictly evaluated and then judged whether they need to be transferred to the ICU for treatment and care, even for emergency cerebral surgery^[16-21]^. Identifying postoperative patients requiring ICU admission is a challenging but necessary daily task. In the clinical setting, some patients undergoing cerebral surgery require unplanned ICU transfer despite thorough evaluation^[22, 23]^. Therefore, it is vital to improve the positive rate of identification, especially for those patients assessed as transferred to the ward, but transferred to the ICU after surgery.

The identification of risk factors is critical to improving preoperative assessment. Therefore, this study aimed to examine preoperative, surgical, and PACU data for potential risk factors for unplanned ICU transfer, which has implications for improving the management and postoperative resource allocation of those vulnerable patients.

## Materials and methods

### Design

This was an observational, retrospective, case-control study in which each case was matched with two controls. Unplanned transfer to the ICU was defined as no plan to transfer the patient to the ICU before starting anesthesia (i.e., the decision to transfer to ICU was made during or after surgery).

### Setting

The study was conducted at the Affiliated Hospital of Jining Medical University, a Shandong Provincial Regional Medical Center with 82 clinical departments, nine intensive care units, and 3,028 beds. We retrospectively reviewed data of patients admitted to PACU after cerebral surgery. Those scheduled to be transferred to the ICU were excluded. Because for patients with scheduled postoperative intensive care admission, the general practice is to transfer them directly from the operating room to the ICU. The study followed the guidelines of the Declaration of Helsinki. Ethics committee/IRB of Affiliated hospital of Jining Medical University gave ethical approval for this work (approval number: 2021-07-C009). A written informed consent was obtained from all patients upon transfer to the Affiliated Hospital of Jining Medical University.

### Cases

Electronic records were used to selected patients ≥18 years of age who had undergone cerebral surgery from January 1, 2016 to December 31, 2020 and who had an unplanned transfer to ICU after cerebral surgery. The researchers reviewed the completeness and accuracy of each case.

### Controls

For each patient with an unplanned transfer to ICU after cerebral surgery, two controls of the same type of surgery were randomly selected from the electronic medical records. Patients in the control group were matched (2:1) for sex, age (±5 years), and operation date (±48 hours). Researchers reviewed the data from patients in the control group to ensure the integrity of the included patients’ medical records.

### Data collection

All data extraction and data input were completed by members of the research team who have been trained in scientific research.

The following demographic and disease history data were collected: age, sex, smoking history, drinking history, and the presence of lung disease, hypertension, diabetes, hyperlipidemia, and/or cardiovascular disease. Patients who had quit smoking and drinking were still categorized into the smoking and drinking groups, respectively. Data on laboratory indicators, such as platelet volume, hemoglobin, total protein, albumin, uric acid, and blood glucose levels, were also collected. Laboratory indicators were measured on an empty stomach the day before the operation.

Surgical and anesthetic data, including the type of surgery, operation duration, blood loss volume, fluid replacement volume, urine volume, blood transfusion volume, heart rate, percentage of oxygen saturation (SpO_2_), removal of tracheal intubation, reintubation, and PACU duration, were collected. Volume of blood loss, of fluid replacement, and of urine were obtained over the entire operation period; heart rate and blood oxygen were obtained immediately after leaving the operating room.

### Statistical analysis

Statistical analysis was performed using Stata version 14.0 (StataCorp, College Station, TX, USA). Categorical data are described in terms of frequencies and percentages. The normality of continuous data was evaluated using the Kolmogorov–Smirnoff test. Normally distributed data are described in terms of means±standard deviation, and non-normally distributed data in terms of medians and interquartile ranges. The T-test, Mann-Whitney test and the Chi-squared test were used to compare the characteristics between the case and the control groups. Logistic regression analysis was used to identify independent risk factors for unplanned ICU transfer of cerebral surgery patients. Statistical significance was set at p<0.05.

## Results

### Patient characteristics

From January 2016 to December 2020, 11,807 patients received recovery care after cerebral surgery in the PACU, of which 81 patients had unplanned ICU transfer (incidence of unplanned ICU transfer, 0.686%). Among unplanned ICU transfer patients, three patients under the age of 18 years and two patients who lacked relevant data were excluded. Finally, 228 patients were enrolled in this study: 76 in the case group and 152 in the control group (Fig 1).

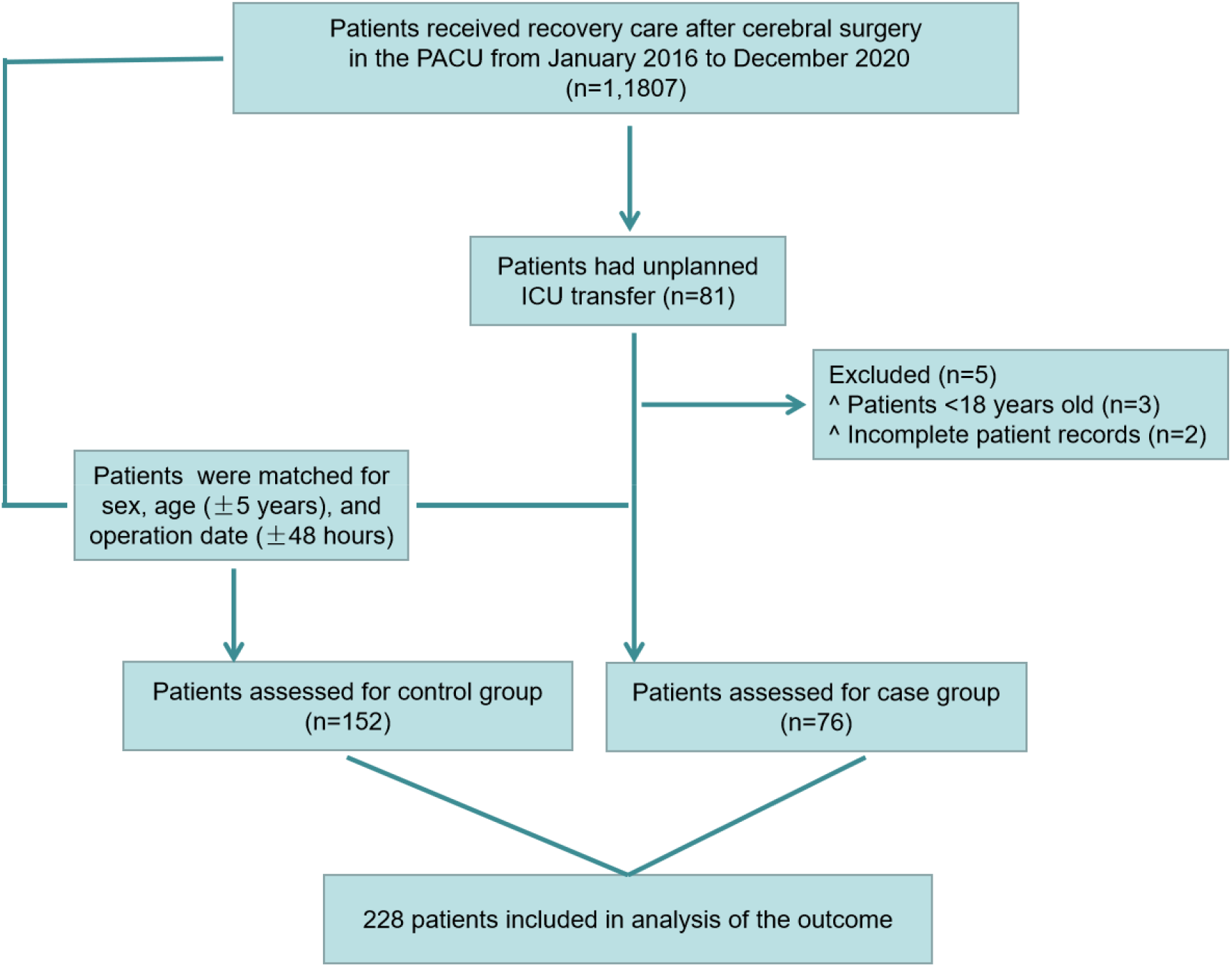

We compared the preoperative, intraoperative, and PACU characteristics of cases receiving routine postoperative care, and those with unplanned ICU transfer. There were a number of significant differences between the two patient populations (Table 1).

**Table 1.**
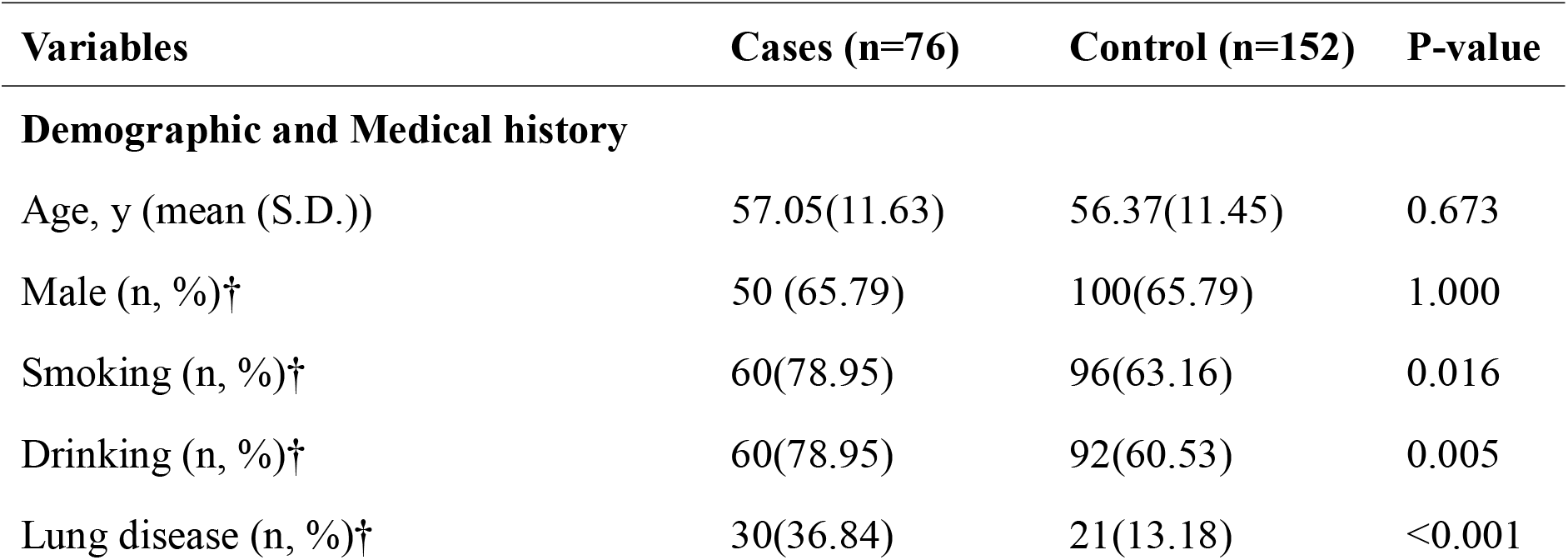
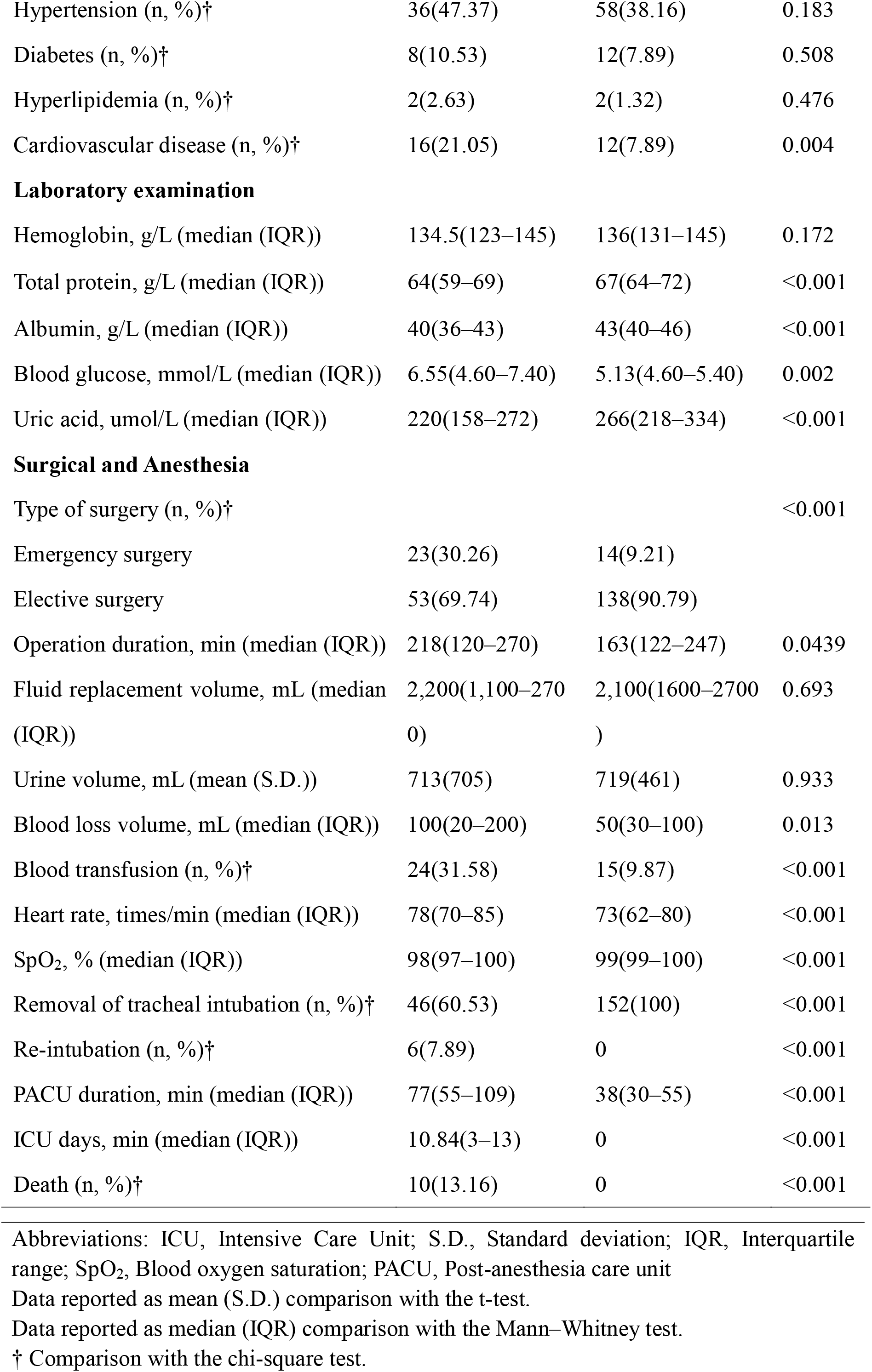
Patient characteristics.

Compared with routine postoperative care, patients who with unplanned ICU transfer were more likely to have a history of smoking and alcohol use; a higher combined rate of pulmonary and cardiovascular disease; lower levels of total protein, albumin, and uric acid; and higher blood glucose levels. They were more likely to undergo emergency surgery, had longer procedures, lost more blood, and receive blood transfusions. They had higher mean heart rates, lower oxygen saturation levels, the possibility of removal of endotracheal tube was lower with higher risk of reintubation, and longer stay in PACU.

### Risk factors for unplanned ICU transfer

In univariate analysis, matching factors were removed, such as age, gender, and consequential factors, including ICU days and number of deaths. We used a simple logistic regression model to analyze the risk factors that may affect postoperative unplanned ICU transfer of patients with cerebral surgery (Fig 2) (Table 2).

**Table 2.** Univariate analysis of unplanned ICU transfer.

| Variables | Odds Ratio (95% CI) | P-value |
| --- | --- | --- |
| Smoking | 2.18 (1.15–4.16) | 0.017 |
| Drinking | 2.45 (1.29–4.64) | 0.006 |
| Lung disease | 3.64 (1.89–7.00) | <0.001 |
| Hypertension | 1.46 (0.83–2.54) | 0.184 |
| Diabetes | 1.37 (0.54–3.51) | 0.509 |
| Hyperlipidemia | 2.03 (0.28–14.68) | 0.484 |
| Cardiovascular disease | 3.11 (1.39–6.97) | 0.006 |
| Hemoglobin | 1.01 (0.99–1.03) | 0.228 |
| Total protein | 1.06 (1.02–1.11) | 0.002 |
| Albumin | 1.12 (1.06–1.19) | <0.001 |
| Blood glucose | 1.70 (1.32–2.19) | <0.001 |
| Uric acid | 1.01 (1.00–1.01) | <0.001 |
| Emergency surgery | 4.28 (2.05–8.93) | <0.001 |
| Operation duration | 1.00 (0.99–1.00) | 0.099 |
| Fluid replacement volume | 1.00 (0.99–1.00) | 0.974 |
| Urine volume | 1.00 (0.99–1.00) | 0.933 |
| Blood loss volume | 1.00 (1.00–1.00) | 0.004 |
| Blood transfusion | 4.22 (2.05–8.66) | <0.001 |
| Heart rate | 1.04 (1.01–1.07) | <0.001 |
| SpO<sub>2</sub> | 1.55 (1.26–1.90) | <0.001 |

Abbreviations: ICU, Intensive Care Unit; CI, confidence interval; SpO<sub>2</sub>, Blood oxygen saturation

**Fig 2.**
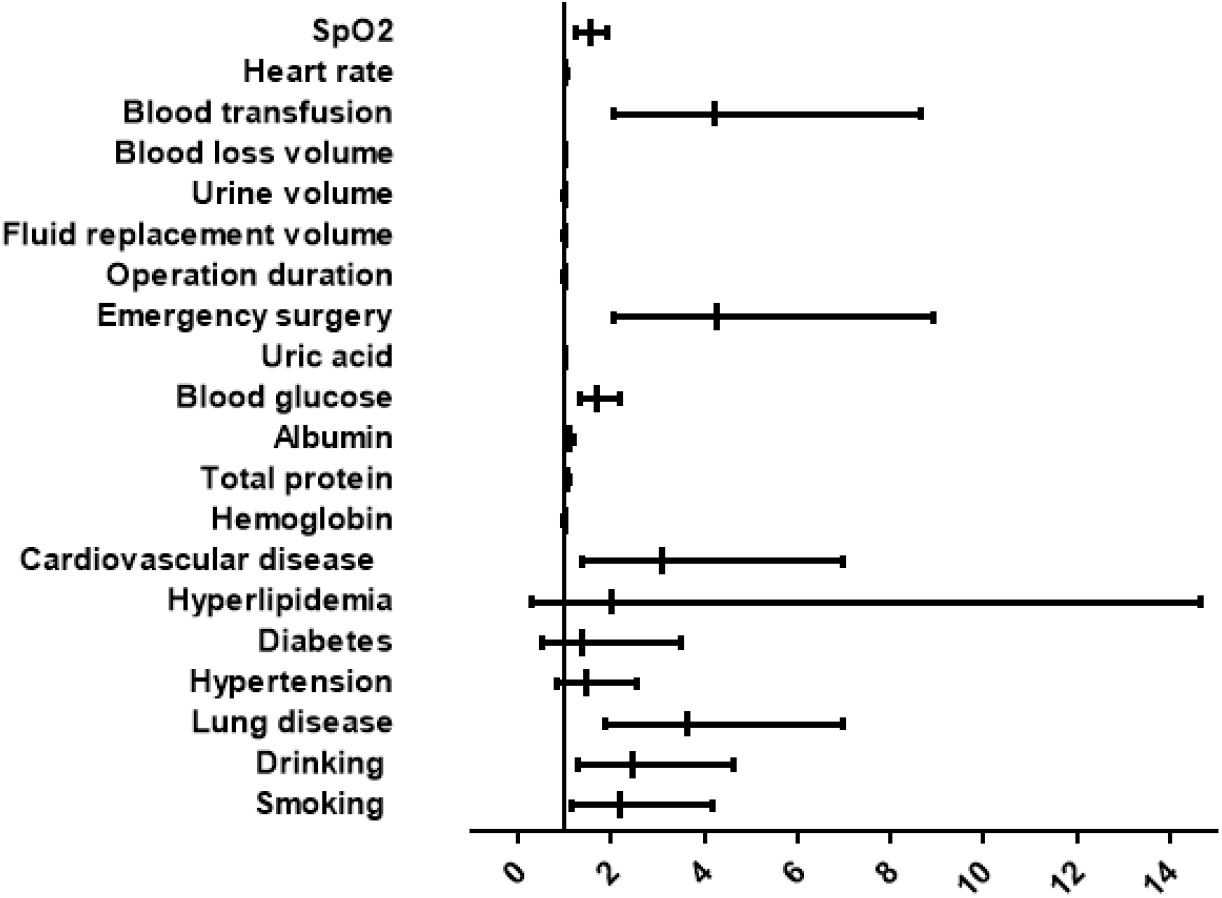
Forest plot for Univariate analysis of unplanned ICU transfers.

All variables at p <0.1 in univariate analysis were further evaluated by multivariate logistic regression to assess independent risk factors for unplanned ICU transfer. Of the initial 20 variables, 7 remained in the final multivariate model (Fig 3) (Table 3).

**Table 3.** Multivariate analysis of unplanned ICU transfer.

| Variables | Odds Ratio (95% CI) | Z | P-value |
| --- | --- | --- | --- |
| Lung disease | 2.64 (1.06–6.60) | 2.08 | 0.037 |
| Albumin | 1.14 (1.03 –1.25) | 2.65 | 0.008 |
| Blood glucose | 1.71 (1.21–2.41) | 3.08 | 0.002 |
| Emergency surgery | 3.08 (1.07–8.87) | 2.09 | 0.037 |
| Blood transfusion | 2.78 (1.02–7.58) | 1.99 | 0.046 |
| Heart rate | 1.04 (1.00–1.08) | 2.01 | 0.045 |
| SpO<sub>2</sub> | 1.57 (1.20–2.04) | 3.32 | 0.001 |

Abbreviations: ICU, Intensive Care Unit; CI, confidence interval; SpO<sub>2</sub>, Blood oxygen saturation

**Fig 3.**
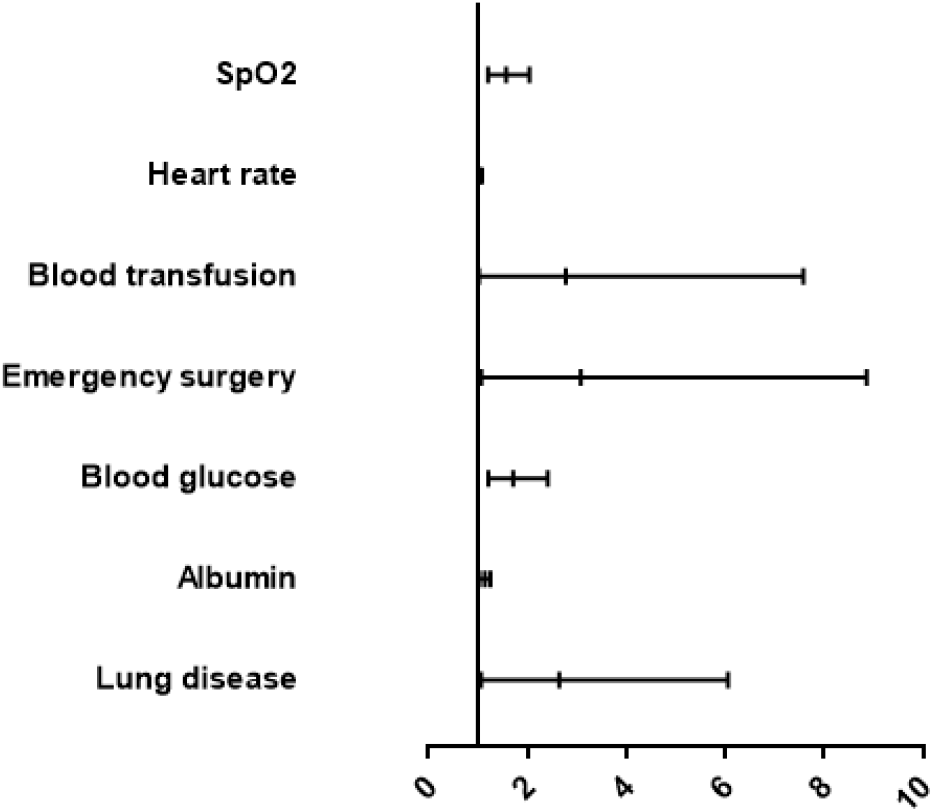
Forest plot for multivariate analysis of unplanned ICU transfers.

## Discussion

This retrospective study aimed to explore the incidence and influencing factors of unplanned ICU transfer from PACU in patients undergoing cerebral surgery. Based on the results of logistic regression, our study found that compared with patients receiving usual care postoperatively, the patients’ characteristics who were transferred from PACU to ICU unplanned after cerebral surgery were those who were more likely to have pulmonary disease, lower mean albumin, higher mean blood glucose, more likely to undergo emergency surgery, more likely to receive intraoperative blood transfusion, higher mean heart rate, and lower mean oxygen saturation. To best of our knowledge, this is the first study to investigate the risk factors for unplanned ICU transfer from the PACU in patients undergoing cerebral surgery. This present studyserves a reference for our next step in developing an electronic medical record system-based, large data-based intraoperative prediction model for unplanned ICU transfer of surgical patients.

We investigated 11,807 patients admitted to the PACU after cerebral surgery and found that 81 had unplanned ICU transfers, with an incidence rate of 0.686%. A study by Hines et al. involved 18,437 patients in the PACU recorded a total of 186 unplanned ICU admissions (1.01%)^[24]^, and a study by Kluger et al. investigated 49,532 patients who were admitted to the PACU for observation and found that 184 had unplanned ICU transfers (0.4%)^[25]^. Overall, the incidence of unplanned ICU transfer in PACU patients ranged from 0.4 to 1.01%. This is similar to the previously reported incidence of unplanned postoperative ICU transfer in adults (0.1–0.9%)^[26-29]^.

Our study showed that 76 patients with unplanned ICU transfer from the PACU had an average ICU length of stay of 10.84 days and an in-hospital mortality rate of 13.16%. A study by Mou et al. found that 40 patients who were unplanned transferred to the ICU by postoperative intermediate care units had an average ICU length of stay of 2.5 days and an in-hospital mortality rate of 12.5%^[30]^. Wanderer et al. reported that the average length of ICU stay was 5 days and the in-hospital mortality rate was 5.6% in 4,847 patients who were unplanned transferred to the ICU from the PACU^[31]^. In contrast, the mean ICU length of stay for patients who were unplanned to be transferred to the ICU in our hospital was slightly higher than that in the other two studies, and the in-hospital mortality rate was similar to that in a certain study. This may be related to the different types of procedures that patients experience.

Of the 76 patients not scheduled for ICU transfer, 6 required emergency intubation (7.89%), corresponding to 0.05% of all patients during the study. Rose et al. found that of 21,457 patients under general anesthesia, 0.1% (n = 22) required emergency reintubation in the PACU^[32]^. Bruins et al. reported that the emergency intubation rate in the PACU of their hospital was also 0.05% (25 out of 49,532 procedures)^[33]^. Peskett et al. found that patients required reintubation was 13 (0.08%) (11 out of 266 procedures) postoperatively^[34]^. In summary, the reintubation rate in PACU centers is approximately between 0.05 and 0.08.

### Clinical and research implications

Unplanned postoperative ICU transfer is worse than planned postoperative ICU transfer for both patients and medical staff. Spontaneous ICU transfer after surgery challenges medical staff to adjust the allocation of ICU resources and psychologically burdens patients and their families. Furthermore, unplanned transfer to ICU usually lead to longer hospital stay, higher medical bills, and worse outcomes^[35, 36]^. Spontaneous ICU transfer also decrease the trust of patients and their families in medical staff^[37]^.

Exploring the risk factors of unplanned transfer to ICU is of great significance to improve the awareness of both medical staff and patients and help them take preventive measures for potential risk factors. In the future, we hope to preoperatively stratify patients based on these factors to predict which patients will need a higher level of postoperative care. By preventing or predicting the need for unplanned transfer to ICU after surgery, we can better allocate ICU resources, reduce the morbidity and mortality of vulnerable patients, and ultimately improve the overall safety of cerebral procedures.

### Strengths and limitations

The study had limitations. First, this study was retrospective in nature; hence, all data collected were from cases and medical records. Second, the complete outcome of the patients was unclear, and further longitudinal studies may be needed. Third, was a single-centered study, and that the results may have been affected by regional characteristics, hospital policies, and types of surgical procedures provided by the hospital. Finally, this study aimed to explore risk factors; thus, the results might have been affected by the included variables.

## Conclusion

The study identified independent risk factors for unplanned transfer from PACU to ICU after cerebral surgery by screening demographic data, laboratory tests, surgical and anesthesia-related information of patients undergoing cerebral surgery. Combining the results of the study with clinical practice in PACU can identify the possibility of unplanned ICU transfer from cerebral surgery patients who do not have an ICU appointment, providing guidance for accurately identifying the patient’s level of care.

## Supporting information

STROBE-checklist-v4-case-control

## Data Availability

The data on which the results presented in the study are based are available from the electronic medical record system of the Affiliated Hospital of Jining Medical University.

## Acknowledgements

We would like to than Yufen Fei, Tong Shen, and Shouxin Zhang for their help in obtaining data and entry for this article. In addition, we are grateful to our colleagues from the Department of Anesthesiology in the Affiliated Hospital of Jining Medical University for their support of the research.

## Notes

**Author Disclosure Statements** All authors indicate that the study does not have any conflicts of interest.

**Funding statement** The research was supported by the Jining Key Research and Development Program (Social People’s Livelihood) project (study code: 2020 YXNS003) hosted by the head nurse Meng Haihong and the “Miao Pu” program of the Affiliated Hospital of Jining Medical University (study code: MP-MS-2020-016) hosted by Cao Qinqin.

### Competing Interest Statement

The authors have declared no competing interest.

### Funding Statement

This study were funded by Jining Key Research and Development Program (Social People's Livelihood) project (study code: 2020 YXNS003) hosted by the head nurse Meng Haihong and the "Miao Pu" program of the Affiliated Hospital of Jining Medical University (study code: MP-MS-2020-016) hosted by Cao Qinqin.

### Author Declarations

Ethics committee/IRB of Affiliated hospital of Jining Medical University gave ethical approval for this work

## Reference

[1] Waddle J P, Evers A S, Piccirillo J F. Postanesthesia care unit length of stay: quantifying and assessing dependent factors[J]. Anesth Analg, 1998, 87(3): 628–633.

[2] Truong L, Moran J L, Blum P. Post anaesthesia care unit discharge: a clinical scoring system versus traditional time-based criteria[J]. Anaesth Intensive Care, 2004, 32(1): 33–42.

[3] Lee L, Tran T, Mayo N E, et al. What does it really mean to “recover” from an operation?[J]. Surgery, 2014, 155(2): 211–216.

[4] Bothner U, Georgieff M, Schwilk B. The impact of minor perioperative anesthesia-related incidents, events, and complications on postanesthesia care unit utilization[J]. Anesth Analg, 1999, 89(2): 506–513.

[5] Bruins S D, Leong P M, Ng S Y. Retrospective review of critical incidents in the post-anaesthesia care unit at a major tertiary hospital[J]. Singapore Med J, 2017, 58(8): 497–501.

[6] Chomton M, Marsac L, Deho A, et al. Transforming a paediatric ICU to an adult ICU for severe Covid-19: lessons learned[J]. Eur J Pediatr, 2021, 180(7): 2319–2323.

[7] Coppadoro A, Benini A, Fruscio R, et al. Helmet CPAP to treat hypoxic pneumonia outside the ICU: an observational study during the COVID-19 outbreak[J]. Crit Care, 2021, 25(1): 80.

[8] Bing-Hua Y U. Delayed admission to intensive care unit for critically surgical patients is associated with increased mortality[J]. Am J Surg, 2014, 208(2): 268–274.

[9] Shank C D, Erickson N J, Miller D W, et al. Reserved Bed Program Reduces Neurosciences Intensive Care Unit Capacity Strain: An Implementation Study[J]. Neurosurgery, 2020, 86(1): 132–138.

[10] Hourmant Y, Mailloux A, Valade S, et al. Impact of early ICU admission on outcome of critically ill and critically ill cancer patients: A systematic review and meta-analysis[J]. J Crit Care, 2021, 61: 82–88.

[11] Zhou Y T, Tong D M, Wang S D, et al. Acute spontaneous intracerebral hemorrhage and traumatic brain injury are the most common causes of critical illness in the ICU and have high early mortality[J]. BMC Neurol, 2018, 18(1): 127.

[12] Franko L R, Hollon T, Linzey J, et al. Clinical Factors Associated With ICU-Specific Care Following Supratentoral Brain Tumor Resection and Validation of a Risk Prediction Score[J]. Crit Care Med, 2018, 46(8): 1302–1308.

[13] de Almeida C C, Boone M D, Laviv Y, et al. The Utility of Routine Intensive Care Admission for Patients Undergoing Intracranial Neurosurgical Procedures: A Systematic Review[J]. Neurocrit Care, 2018, 28(1): 35–42.

[14] Ziai W C, Varelas P N, Zeger S L, et al. Neurologic intensive care resource use after brain tumor surgery: an analysis of indications and alternative strategies[J]. Crit Care Med, 2003, 31(12): 2782–2787.

[15] Ehlers L D, Pistone T, Haller S J, et al. Perioperative risk factors associated with ICU intervention following select neurosurgical procedures[J]. Clin Neurol Neurosurg, 2020, 192: 105716.

[16] Bonow R H, Quistberg A, Rivara F P, et al. Intensive Care Unit Admission Patterns for Mild Traumatic Brain Injury in the USA[J]. Neurocrit Care, 2019, 30(1): 157–170.

[17] Volovici V, Ercole A, Citerio G, et al. Intensive care admission criteria for traumatic brain injury patients across Europe[J]. J Crit Care, 2019, 49: 158–161.

[18] Taylor W A, Thomas N W, Wellings J A, et al. Timing of postoperative intracranial hematoma development and implications for the best use of neurosurgical intensive care[J]. J Neurosurg, 1995, 82(1): 48–50.

[19] Lonjaret L, Guyonnet M, Berard E, et al. Postoperative complications after craniotomy for brain tumor surgery[J]. Anaesth Crit Care Pain Med, 2017, 36(4): 213–218.

[20] Henker C, Schmelter C, Piek J. [Complications and monitoring standards after elective craniotomy in Germany][J]. Anaesthesist, 2017, 66(6): 412–421.

[21] Laan M T, Roelofs S, Van Huet I, et al. Selective Intensive Care Unit Admission After Adult Supratentorial Tumor Craniotomy: Complications, Length of Stay, and Costs[J]. Neurosurgery, 2020, 86(1): E54–E59.

[22] Bui J Q, Mendis R L, van Gelder J M, et al. Is postoperative intensive care unit admission a prerequisite for elective craniotomy?[J]. J Neurosurg, 2011, 115(6): 1236–1241.

[23] Laan M T, Roelofs S, Van Huet I, et al. Selective Intensive Care Unit Admission After Adult Supratentorial Tumor Craniotomy: Complications, Length of Stay, and Costs[J]. Neurosurgery, 2020, 86(1): E54–E59.

[24] Hines R, Barash P G, Watrous G, et al. Complications occurring in the postanesthesia care unit: a survey[J]. Anesth Analg, 1992, 74(4): 503–509.

[25] Kluger M T, Bullock M F. Recovery room incidents: a review of 419 reports from the Anaesthetic Incident Monitoring Study (AIMS)[J]. Anaesthesia, 2002, 57(11): 1060–1066.

[26] Meziane M, El J S, Elkoundi A, et al. Unplanned Intensive Care Unit Admission following Elective Surgical Adverse Events: Incidence, Patient Characteristics, Preventability, and Outcome[J]. Indian J Crit Care Med, 2017, 21(3): 127–130.

[27] Da S P,de Aguiar V E, Fonseca M C. Risk factors and outcomes of unplanned PICU postoperative admissions: a nested case-control study[J]. Pediatr Crit Care Med, 2013, 14(4): 420–428.

[28] Petersen T M, Ludbrook G L, Flabouris A, et al. Developing models to predict early postoperative patient deterioration and adverse events[J]. ANZ J Surg, 2017, 87(6): 457–461.

[29] Petersen T M, Ludbrook G L, Flabouris A, et al. Developing models to predict early postoperative patient deterioration and adverse events[J]. ANZ J Surg, 2017, 87(6): 457–461.

[30] Fujii T, Uchino S, Takinami M. Life-threatening complications after postoperative intermediate care unit discharge: A retrospective, observational study[J]. Eur J Anaesthesiol, 2016, 33(1): 22–27.

[31] Wanderer J P, Anderson-Dam J, Levine W, et al. Development and validation of an intraoperative predictive model for unplanned postoperative intensive care[J]. Anesthesiology, 2013, 119(3): 516–524.

[32] Rose D K. Recovery room problems or problems in the PACU[J]. Can J Anaesth, 1996, 43(5 Pt 2): R116–R128.

[33] Bruins S D, Leong P M, Ng S Y. Retrospective review of critical incidents in the post-anaesthesia care unit at a major tertiary hospital[J]. Singapore Med J, 2017, 58(8): 497–501.

[34] Peskett M J. Clinical indicators and other complications in the recovery room or postanaesthetic care unit[J]. Anaesthesia, 1999, 54(12): 1143–1149.

[35] Liu V, Kipnis P, Rizk N W, et al. Adverse outcomes associated with delayed intensive care unit transfers in an integrated healthcare system[J]. J Hosp Med, 2012, 7(3): 224–230.

[36] Gold C A, Mayer S A, Lennihan L, et al. Unplanned Transfers from Hospital Wards to the Neurological Intensive Care Unit[J]. Neurocrit Care, 2015, 23(2): 159–165.

[37] Jennerich A L, Hobler M R, Sharma R K, et al. Unplanned Admission to the ICU: A Qualitative Study Examining Family Member Experiences[J]. Chest, 2020, 158(4): 1482–1489.

